# Basic emergency Obstetric and Neonatal Care Knowledge retention and Skills of Health Professionals in Burundi following an ALARM International Program Training: A pilot study

**DOI:** 10.1101/2020.05.11.20098632

**Authors:** Anifa Luyinga Kalay, Veronique Mareschal, Juma Ndereye, Jocelynn Cook

## Abstract

**Background:** This study describes the immediate and short-term improvement in basic emergency obstetric and neonatal care (BEmONC) knowledge and skills after ALARM International Programme (AIP) training in Burundi.

**Subjects and Method:** In 2017, sixteen health professionals participated in a 5-day AIP training. They completed pre and post course tests. At the end of the training, they did objective structured clinical examinations (OSCE) on postpartum hemorrhage (PPH), shoulder dystocia and neonatal resuscitation. In 2018, a refresher course was offered to participants who scored the highest in the 2017 post-training test. Pre and post tests were administered. Mean pre- and post-test scores and t-paired test was performed to determine the knowledge change and retention between 2017 and 2018. Mean OSCE scores were calculated to describe acquired clinical skills at the 2017 training. A one-way ANOVA test was performed to assess the differences between cadres’ scores in the tests and the OSCEs.

**Results:** In 2017, pre and post scores were significantly different within cadres except imidwives: physicians t(5)=8.77(p<0.05), midwives t(1)=5,29 (p=0,11) and nurses t(7)=7,91(p<0.05) In 2018, the mean score was 67% (± 10.31) on the pre-test and was 82% (± 5.78) in 2017 post-test. The difference was significant (t(7)=4.80, p<0.05). Scores were significantly different between cadres in PPH [F=(2,13)=6.17, p <0.05)] and dystocia [F=(2,13)=3.92, p <0.05)].

**Conclusion:** An immediate gain in knowledge and acquisition of critical skills was observed in all cadres. Knowledge significantly decreased eight months following AIP training. Supervision, mentoring of trainees and continuing medical education are needed following the initial training.

## BACKGROUND

In 2000, United Nations countries set 8 Millenium Development Goals (MDG) to be reached by 2015. Although all goals had the potential to impact on health a potential impact on health, two of them directly pertained to maternal and child health: reducing child mortality (goal 4) and improving maternal health (goal 5) (United Nations, 2015)

Burundi was one of the many African countries that did not meet either of the aforementioned goals. Child mortality did decrease, but not at two-thirds of the 1990 rate as targeted MDG 4). Under five mortality rate was 180 per 1,000 live birth in 1990 and 81 per 1,000 live births in 2017 (Institute of Health Metrics, 2015). About one third of the under 5 mortalities were from death occurring during the first month of life (Gouvernement of Burundi, 2010)

Burundi’s maternal mortality ratio decreased from 1,220 per 100 000 live births in 1990 to 715 per 100 000 live births in 2015. (World Health Organization, 2015) However, this did not represent the three quarter reduction from the 1990 rate as set for MDG 5.

Neonatal mortality is an instrumental issue to address in order to significantly decrease under 5 mortality. Various factors have been identified to explain the slow decrease in neonatal mortality, including limited implementation of essential care for newborns, poor quality of mother and newborn care in the areas of emergency obstetrical and neonatal care (EmONC) and postnatal care. (Gouvernement of Burundi, 2010)

Maternal deaths in Burundi are mainly due to direct obstetrical causes (i.e. postpartum hemorrhage, dystocia and infections (Gouvernement of Burundi, 2010). Poor EmONC quality, limited number and insufficiently trained health professionals as well as the low quality of maternal health services in general have been associated, among others, with the slow reduction in the rate of maternal mortality in Burundi. (Gouvernement of Burundi, 2010)

EmONC quality is an important element to improve maternal and children mortality in Burundi. A recent assessment of Basic and Comprehensive EmONC (BEmONC and CEmONC) conducted in 2010 revealed that only 2% of health centres and hospitals in Burundi had the infrastructure required to be able to deliver BEmONC and CEmONC functions. (Chi *et al*., 2015)

As for human resources in EmONC, both a shortage of health professionals as well as their inadequate training in EmONC have been identified as barriers to the delivery of EmONC in Burundi (Chi *et al*., 2015)

Furthermore, the quality of the EmONC training differed across the country depending on which institution was responsible for delivering the training. The lack of hands-on skills sessions in the majority of the training programs were also identified as deleterious to quality EmONC training.

In 2014, Burundi’s Ministry of Health‘s national Program of reproductive Health developed in-service training programs in BEmONC and CEMONC for nurses, midwives and physicians. Both courses were 6 days in duration, and the CEMONC included an 8-week internship. Various teaching approaches were used; lectures, group discussions, role play, case studies and simulations (UNICEf et al., 2014). However, due to the length of the course and the associated cumbersome logistics, few health professionals have been trained to date. A shorter and efficient in-service training would be more cost-efficient and sustainable in the fragile context of Burundi.

Thus, in a project implemented in Bujumbura by L’AMIE(Aide internationale à l’enfance), the Society of Obstetricians and Gynaecologists of Canada(SOGC) proposed to use the SOGC’s Advances in Labour and Risk Management (ALARM) International Program (AIP) to train health professionals. The purpose of this project was to decrease maternal and neonatal mortality by increasing capacity and support of health care providers’ training in obstetrical care.

The ALARM International Program (AIP) is a five-day training course which covers the main causes of maternal and neonatal death in developing countries, grounded in a sexual and reproductive health and rights approach. Topics such as evidence-based medicine, risk management, maternal death audits and communication are also covered. An optional sixth day of training can be added to train AIP trainers.

The AIP has been used in twenty countries and has been translated into French and Spanish and its effectiveness has been demonstrated. For instance, after AIP training in Mali and Senegal, it was observed that hospital-based maternal mortality was reduced by 15% over a 2 year period; death from hemorrhage, preeclampsia/eclampsia and puerperal infection as well as neonatal mortality before 24 hours decreased (Dumont *et al*., 2013). In Kenya, an evaluation of the impact of the AIP conducted one year after the training showed an improvement in post-partum hemorrhage (PPH) outcomes, use of oxytocin and fewer neonates with 5-minute Apgar scores of less than 5 (Spitzer *et al*., 2014).

The present study describes the immediate and short-term improvement in BEmONC knowledge and skills following a one-time AIP training of health care providers practicing in Bujumbura, Burundi.

## SUBJECTS AND METHODS

The present study was implemented within a larger project aiming at contributing to decreasing the maternal and newborn mortality in Bujumbura, which focused on improving community awareness of maternal and neonatal issues, making maternal and newborn care services available to the population in the targeted area and strengthening health professionals’ capacity (L’AMIE, 2016).

With a population of about 1.2 million (PopulationData.net, 2019). The province of Bujumbura is divided into 9 communes (Wikipedia, 2019). The current study was conducted in the commune of Kanyosha which has a population of approximately 80 000 (Citypopulation.de, 2019), serviced by 22 healthcare facilities; 6 hospitals and 16 health centers. BEmONC is delivered in health centers and CEmONC is available in hospitals. Approximately 200 obstetricians-gynaecologists, general practitioners, midwives and nurses work in these centers and hospitals.

In September 2017, sixteen health professionals were selected to be trained as trainers in BEmONC using the AIP. The main criteria for selection were being a health professional currently working in a health center or hospital in the Kanyosha district and being involved in BEmONC in the last 2 years. Because of security concerns in Burundi, the course was conducted in Kigoma, Tanzania and delivered by 3 SOGC members, one SOGC staff, the director of the National Programme of Reproductive Health and one Burundi Ministry of Health member.

The contents of the AIP was adapted to the needs and existing guidelines of Burundi health professionals in close collaboration with the National Programme of Reproductive Health. For example, contents on forceps use was not covered as they are not used in Burundi. Moreover, the wording was also reviewed to reflect Burundi’s cultural and medical reality. Also, the illustrations used in the lectures were reviewed to ensure cultural adequacy

The AIP training lasted five days and covered 10 topics. Different teaching approaches were used such as lectures, group discussions, role play and hands-on skills activities. Participants were physicians, midwives and nurses. On Day 1, participants completed a pre-course written knowledge test, and wrote a post-training assessment test on the final day. Hands-on skills were also assessed, only at the end of the training, through objective structured clinical examinations (OSCE) on postpartum hemorrhage, shoulder dystocia and neonatal resuscitation.

Each OSCE lasted 9 minutes. Participants read a short written scenario describing the clinical scenario. They were asked to explain the management of the case while demonstrating specific skills on a mannequin. The AIP instructor also asked set questions to assess the participant’s mastery of the most important skills needed to save the mother and the baby.

In May 2018, a refresher course was offered in Kigali, Rwanda to the 8 participants who scored the highest in the post-training tests in September 2017.

A written test was again administered at the beginning and end of the refresher course. This test was the same as the one used in the post-test of the initial training.

The mean pre- and post-test scores was calculated and a t-paired test was performed to determine the extent in knowledge change between pre-test and post-test scores in September 2017 and May 2018. The same statistical computations were performed between the September 2017 post-test scores and the May 2018 pre-test score in order to assess retention. Scores to questions related to PPH and neonatal care were also compared using the same statistical analysis between the September 2017 post test scores and May 2018 pretest scores. These two topics were selected as they are the most important causes of maternal and neonatal deaths in Burundi.

Mean OSCE scores were also calculated to describe acquired clinical skills at the September 2017 training.

A one-way ANOVA test was performed to assess the significance of differences between cadres scores in the written tests and the OSCEs.

This study did not require ethical approval as it was considered as a quality improvement study by the Research Ethics Committee of the Ottawa Hospital.

## RESULTS

### Participants’ profile

In September 2017, 16 health professionals participated in the training. In May 2018, eight health professionals participated in the refresher course. Table 1 shows the characteristics of each group.

**Table 1:** participants profile

| ***Cadres*** | ***September 2017 (n=16)*** |  | ***May 2018 (n=8)*** |  |
| --- | --- | --- | --- | --- |
|  | **Female** | **Male** | **Female** | **Male** |
| Physicians | 2 | 4 | 2 | 4 |
| Midwives | 2 | 0 | 1 | 0 |
| Nurses | 4 | 4 | 0 | 1 |
| TOTAL | 8 | 8 | 3 | 5 |

### Knowledge acquisition

In September 2017, the mean pretest score was 38 % (± 14.06) and the mean post-test score was 73% (± 12.7). The difference was significant (t(15)=11.18 (p<0.05), with the greatest improvement in the physician group. The nurses had the lowest pre and post test scores but still showed significant improvement. Figure 1 shows the differences between groups.

**Figure 1.**
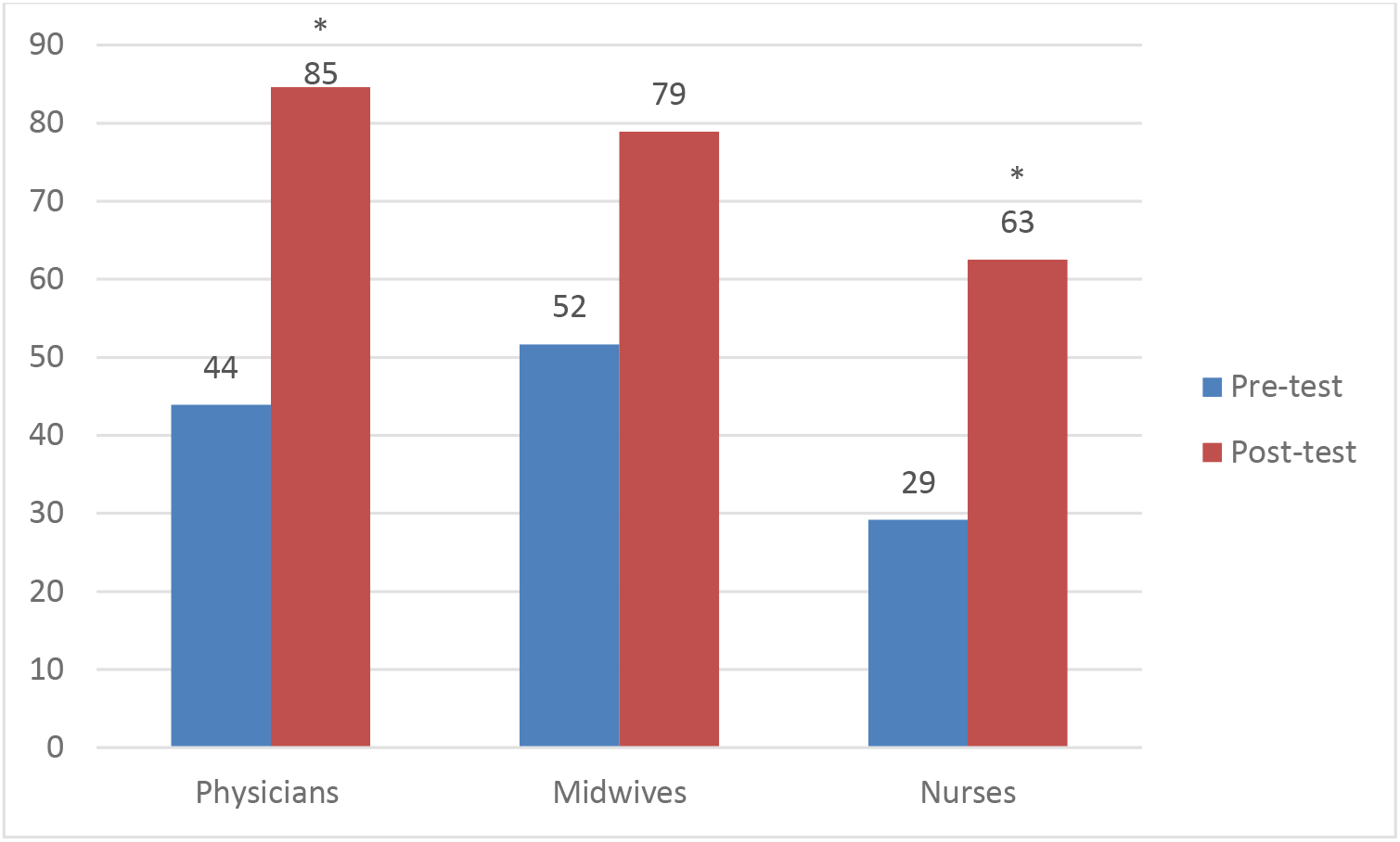
Pre and post-test scores per cadre.

The pre and post scores were significantly different within groups except in midwives: physicians t(5)=8.77(p<0.05), midwives t(1)=5,29 (p=0,11) and nurses t(7)=7,91(p<0.05)

The scores were significantly different between the 3 cadres in the pretest[F=(2,13)=3.97, p <0.05)] and post-scores [F=(2,13)=17.24, p <0.05)]. A Bonferonni corrected t-test indicated that physicians had a significantly higher scores that nurses in the post test [(t (12)=5,61 (p <0.05)]] The other comparisons did not yield significant differences.

### Knowledge retention

In May 2018, the group of eight participants participating in the refresher course obtained a mean score of 67% (± 10.31) on the pre-test whereas their mean September 2017 post-test score was 82% (± 5.78). The difference was significant (t(7)=4.80, p<0.05).Figure 2 shows the individual variation between September 2017 and May 2018. All, but one (#4) participants had lower scores in May 2018.

**Figure 2.**
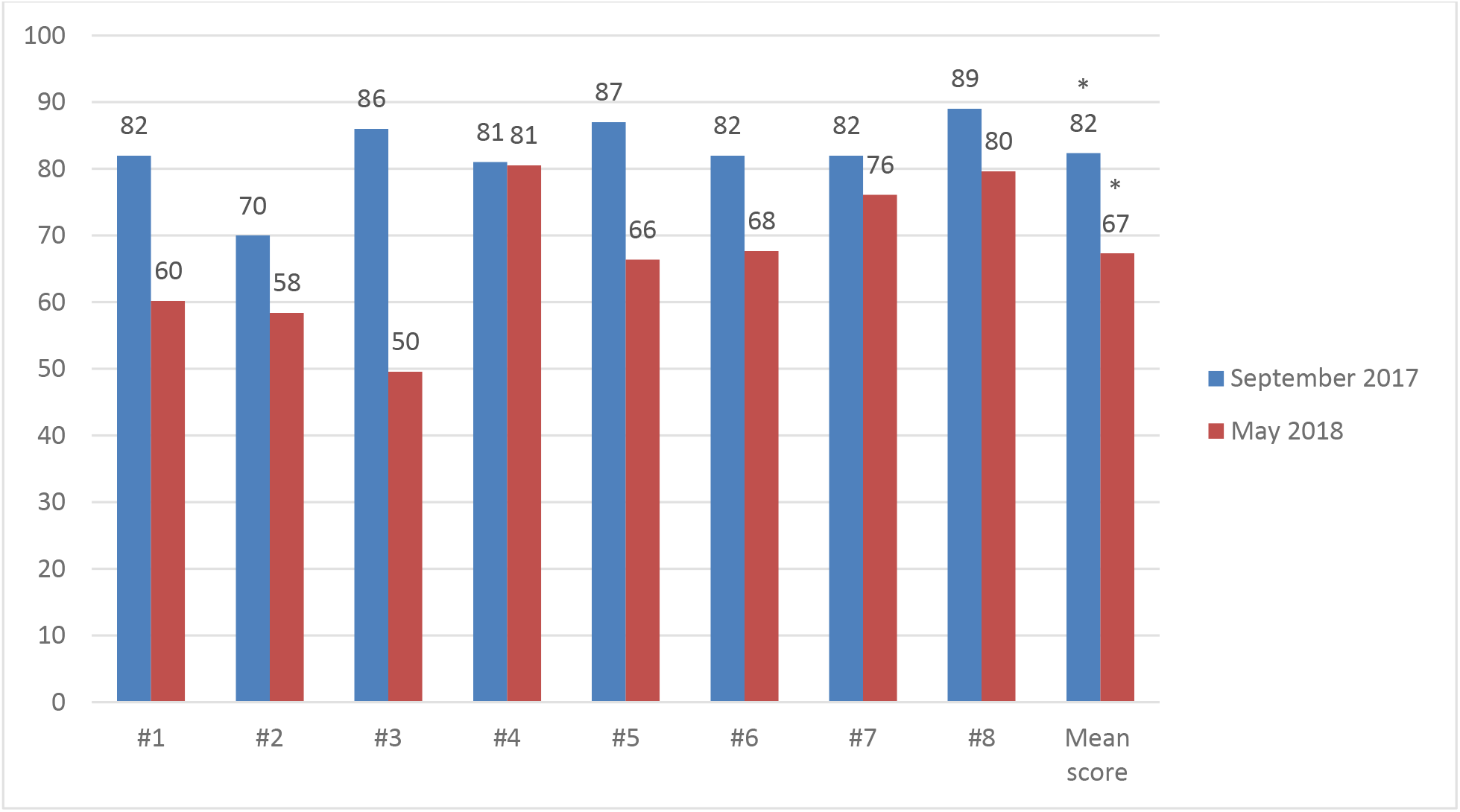
Individual scores variation between September 2017 and May 2018.

### BEmONC skills

With regard to the OSCE administered only at the end of the training in September 2017, the highest scores were observed in the PPH OSCE and the lowest in the neonatal resuscitation one. Nurses performed the lowest in all OSCEs and physicians had the highest scores. Figure 3 shows the OSCE scores per cadre.

**Figure 3.**
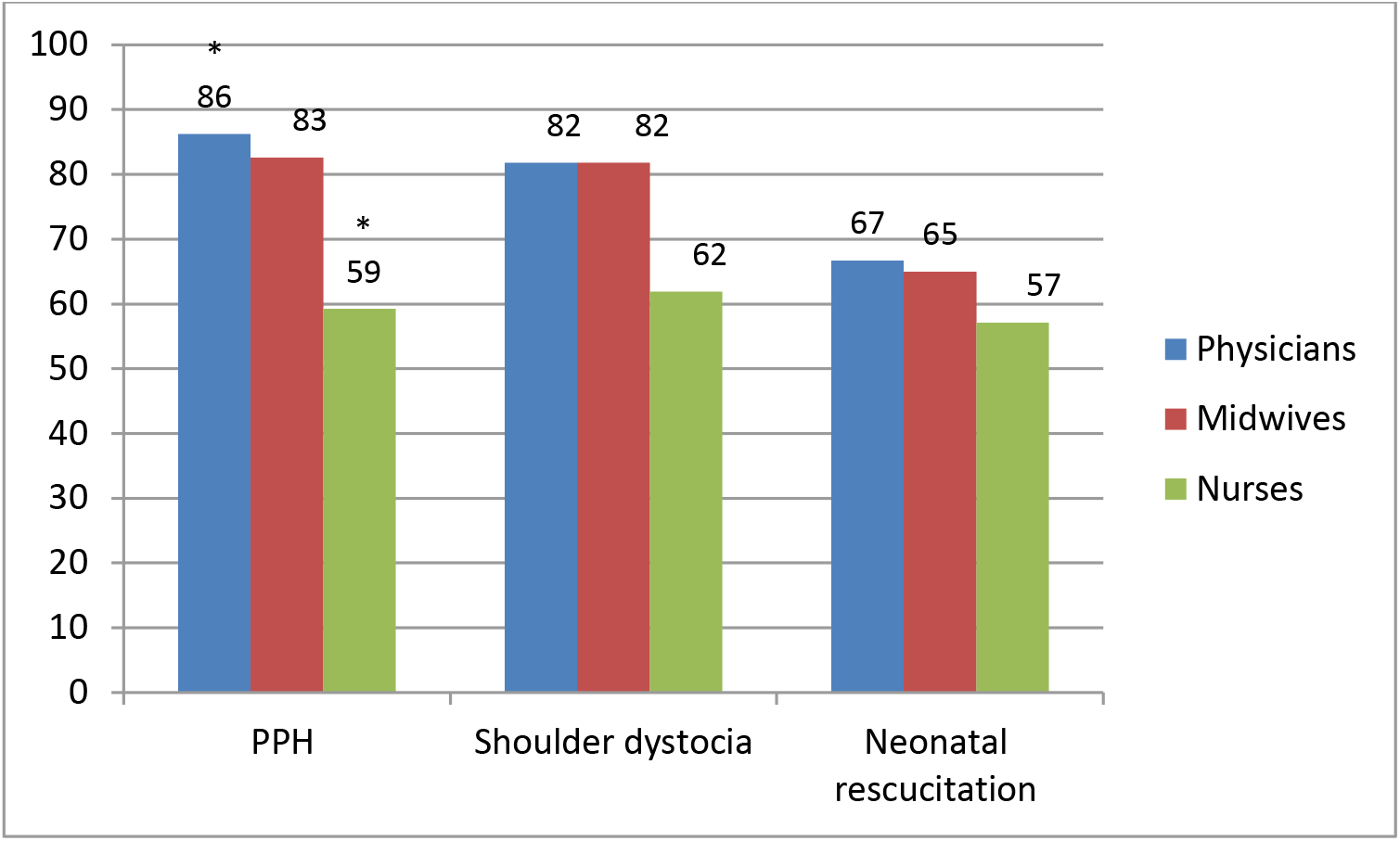
September OSCE scores per cadre.

The scores were significantly different between the 3 cadres in the two OSCEs PPH [F=(2,13)=6.17, p <0.05)] and the dystocia PPH [F=(2,13)=3.92, p <0.05)] The Bonferonni corrected t-test showed that physicians had significantly higher scores that nurses did in the PPH OSCE [(t(12)=-3,22 (p <0.05)]]. The other comparisons were not significant.

## DISCUSSION

A number of BEmONC training programs have been developed by different organizations in the last 25 years (Ameh et al, 2015). The AIP is unique in its design, covering not only the main causes of maternal and newborn deaths in developing countries but also several other areas that impact outcomes such as other obstetrical conditions, evidence-based approaches, maternal death audits and sexual and reproductive health and rights (SRHR). To our knowledge, the AIP is the only BEmONC training that integrates SRHR in all aspects of the teaching and presents a workshop dedicated to this topic. The extent of the AIP content and the short duration of the course makes this training appropriate for limited resource settings for in-service training. Although the longer-term associations of the AIP to improving maternal-neonatal outcomes have been demonstrated, this is the first published study demonstrating the immediate and shorter-term impact of the training on knowledge and skills acquisition and retention in different cadres. This study may guide decision makers in determining not only training needs of each cadre but also the ideal timespan between trainings in order to maintain providers’ knowledge and skills.

Although the sample size was low, the results of this study still demonstrates that, at the end of the training, all professional cadres significantly improved their level of BEmONC knowledge. Most importantly, it is to be noted that the pretest scores were low, suggesting that all participants may not have had similar prior exposure to the range of clinical topics included in the AIP training curriculum. This is reasonable, given that there are limited opportunities for nurses and midwives, in their practice, to perform some clinical procedures (i.e., vacuum extraction or manual vacuum aspiration), and they simply may not have had the training to be knowledgeable or the exposure to be proficient. The same situation applies to physicians who are working in health centers and do not manage high risk pregnancies. Moreover opportunities for continuing medical education are scarce in developing countries and even rarer in fragile countries such as Burundi.

Another observation was that nurses’ scores, although improved, stayed much lower than the other cadres’ especially when compared to physicians. Clearly, given the critical role that nurses play in maternal and newborn health and care, there is an urgent need to explore the underlying reasons for this discrepancy. In other countries, the content and the delivery of the AIP material was adapted to fit the needs of specific cadres such as traditional birth attendants in Mali or medical assistants in Malawi, based on the recognition that literacy and/or technical competency may not be equivalent to the other professional groups. This approach was effective in improving knowledge and skills of the participants (unpublished). Future studies should identify specific needs in the different cadres and adapt the training material to focus on these areas.

Another issue to consider when training different professional groups: the AIP design is based on a multidisciplinary approach in learning by providing training to different cadres at the same time, based on the effectiveness of training “real-life” scenarios where individuals work together in clinical situations. That said, this approach may need to be adapted according to the existing level of knowledge and skills mastery of participants. In Canada, participants in the ALARM program have to read the material and pass the knowledge test before attending the actual training. This approach could be adapted for developing countries in order to identify participants’ strengths and weaknesses before the start of the training. The instructors would then be able to alter their teaching and the clinical emphasis accordingly.

In this particular experience, participants’ knowledge decreased significantly but was still above average 8-months following the AIP training. Several factors can influence knowledge retention including the lack of opportunities to practice acquired knowledge and skills (Tang *et al*., 2016), the type of facility where the participant is working, professional qualification (Mzurikwao, Ng’weshemi Kapalata and Ernest, 2018)and work load (London School of Topical Medicine). Future studies should explore the link between these factors and knowledge retention when using the AIP.

Our findings are similar to previous studies that describe a decrease in knowledge after 9 months. (Ameh *et al*., 2018) This data supports the need for continuing medical education for perinatal care providers of all cadres in Burundi and suggests that the introduction of novel approaches is warranted. Continuous mentoring is an effective tool to help providers to maintain their skills, and this could be performed remotely. It would also encourage communication between local health care providers. The SOGC instructors in this project set up an informal mentoring system using Whatsapp. This mentoring system linked SOGC instructors to Burundese trainees after the AIP training in order to provide them continuous support through discussion of cases encountered in their practice and sharing of relevant information on BEmONC. To date, this is working well and the group is active.

With regard to post training skill assessment, the lowest scores were reported in neonatal resuscitation for all cadres. This skill is an important one to focus on in the hopes of decreasing neonatal death rates in developing countries. Several reasons can explain the poor performance: some participants had had limited to no training on this topic, few opportunities were available to resuscitate a baby especially for those working in health centers, and participants were not familiar with the equipment as they lacked it in their facility.

This study had several limitations. The sample size was small making the generalization of the results difficult, and studies involving a larger sample are needed. Another limit is the lack of formal assessment of skills before the training. However, post training skills scores did reveal the need for more training in neonatal resuscitation which is a known actual concern in Burundi. Factors that influence knowledge retention and skills acquisition were not collected. This study used routine data collected during AIP training. Future research should identify the factors that need to be targeted to improve training outcomes.

### Conclusion

The AIP is an effective tool to improve BEmONC capacity in resource-limited settings. In the current study, a significant immediate gain in knowledge and acquisition of critical skills for management of common issues in BEmONC was observed in all cadres. However, data also revealed that knowledge significantly decreased eight months following AIP training. This shows the need for supervision and mentoring of trainees as well as continuing medical education following the initial training.

## Data Availability

Data are available from the authors on request.

## ACKNOWLEDGEMENTS

The authors thank Chantal Raymond and Janet Northcott, SOGC members, for their technical assistance as well as Denise Bantegeyeko, project coordinator at the APECOS, Burundi for the logistic support through the organization of the courses.

